# Assessing the knowledge and practices of primary healthcare workers on malaria diagnosis and related challenges in view of COVID-19 outbreak in a Nigerian Southwestern Metropolis

**DOI:** 10.1101/2022.05.20.22275356

**Authors:** Esther Ayandipo, Deborah Babatunde, Oladipo Afolayan, Olabisi Kalejaye, Taiwo Obembe

**Affiliations:** Management Sciences for Health; APIN Public Health Initiatives; University College Hospital, Ibadan; Learning & Development Bureau; Claremont Graduate University, California USA

**Keywords:** Malaria, COVID-19, Malaria RDT, Primary Healthcare workers

## Abstract

**Background:** The clinical features of COVID-19 and malaria are interrelated. Due to the similarity of symptoms between the two disease states, patients can be incorrectly diagnosed with the other ailment in areas with limited health resources. There is a dearth of knowledge of co-infection between COVID-19 and malaria from healthcare providers’ perspective. Hence, this study assessed the ability of primary healthcare workers to diagnose malaria infection correctly from COVID-19 infection.

**Methods:** A multistage sampling technique was used to select health care workers who were directly involved in malaria case management at 261 government-owned primary health facilities in Oyo State. Socio-demographic characteristics of respondents, knowledge & practices, COVID-19 differential diagnosis and challenges that healthcare workers face regarding malaria diagnosis were obtained using a standardized electronic structured questionnaire. Descriptive statistics, bivariate and multivariate analysis were conducted on data collected and significant results were interpreted at a 5% level of significance.

**Results:** A good percentage of the respondents (81.6%, 74.3%) had good knowledge about malaria and COVID-19 infection. However, the knowledge gained did not translate to practice, as majority (86.2%) of respondents had poor malaria diagnosis practices. Practices relating to COVID-19 differential diagnosis in 69.7% of respondents were also poor. Most of the respondents attributed poor practices to the unavailability of Malaria Rapid Diagnostic Test (mRDT), inadequate training and continuous capacity improvement. Only 12.3% of the respondents had any form of training on malaria diagnosis and treatment in the last five years.

**Conclusion:** Harmonization of regular trainings and continuous on-the job capacity building is essential to improve case identification, diagnosis and management of both ailments. Also, uninterrupted supplies of essential commodities such as mRDT in laboratories will reduce missed opportunities for malaria diagnosis.

## Background

Malaria is a leading cause of death all over the world, and the most common reason for hospital admissions in many African countries (1). Death rates are known to reduce when diagnosed and treated promptly (2). Differentiating malaria from other tropical infections based on patients’ signs and symptoms is becoming increasingly difficult, thus the need for confirmatory laboratory tests (3). Microscopic examination of thick and thin blood smears remains the gold standard of malaria diagnosis. Antigen detection with rapid diagnostic test kits (RDTs) have been found to provide advantages in remote settings where trained laboratory scientists on malaria microscopy are limited (2). As such, in resource-limited areas, clinical diagnosis and empirical judgement is often a common practice among medical practitioners (3).

An outbreak of pneumonia of unknown origin was reported in Wuhan, Hubei Province, China, in December of 2019, with a global spread of severe acute respiratory syndrome coronavirus 2 (SARS-CoV-2) caused by coronavirus disease (COVID-19) (4). The World Health Organization (WHO) declared a pandemic in March 2020, and since then till date, high COVID-19 death rates have been recorded worldwide (4). COVID-19 symptoms vary from person to person in terms of severity, while some individuals remain asymptomatic (5). Similarities between malaria and COVID-19 has been established with both having generic symptoms making immediate diagnosis difficult to establish sometimes (6).

Diagnosing COVID-19 involves the conduct of either a molecular test – reverse transcriptase polymerase chain reaction (RT-PCR) which is the gold standard (7) on upper and lower respiratory tracts, antigen tests and antibody tests (6). The main symptoms for COVID-19 are fever, cough, difficulty in breathing with patients presenting with non-specific symptoms similar to upper respiratory tract infection (5). Patients with malaria more often present with fever, headache, chills, vomiting and myalgia (3). Symptoms of malaria tend to overlap with symptoms of other infectious diseases including COVID-19 (8), thus limiting the ability of clinicians to diagnose without laboratory tests (3). Access to specific laboratory diagnosis techniques is limited in Africa thus promoting clinical diagnosis for malaria (9), allowing the indiscriminate use of antimalarials and reducing the quality of care, especially in resource limited settings The combination of COVID-19 and malaria epidemics has been found to be devastating especially in low- and middle-income countries (LMICs), with suboptimal healthcare systems plagued with inadequate human and financial resources, and weak infrastructures (6).

Even before the outbreak of COVID-19, over diagnosis of malaria has long been an issue (1,10). In a research conducted in Tanzania, many children and adults were treated for malaria even with no evidence of malaria parasites isolated on research slides (1). With the advent of COVID-19 outbreak, a paradigm shift has led to a revision of management protocols of generic symptoms. Due to the peculiarity that both malaria and COVID-19 infection can exist concurrently in Africa, it has been advised that diagnosis for both COVID-19 and malaria be conducted simultaneously for all cases that present with fever to eliminate the possibilities of misdiagnosis and mistreatments especially when co-infection is suspected (8,11). In order that cases of co-infection with malaria and COVID-19 are adequately diagnosed and properly managed, it is important for frontline health workers to be knowledgeable on clinical signs and symptoms of both infections particularly in low-resource settings where gold-standard equipment might be unavailable or inadequate in quantity. This study sought to determine the ability of primary healthcare workers to correctly diagnose malaria and differentiate between malaria and COVID-19 infections in selected primary health care facilities of a Southwestern Nigerian province.

## Methods

This study utilized a cross-sectional study design. Oyo State, with Ibadan as its state capital, is located in South west, Nigeria. Oyo state is situated in the southwest geo-political zone of Nigeria with a population of approximately 6,190,000 spread over an estimated land mass of about 28,000.00 square kilometres. Oyo State consists of 33 local government areas (LGAs) that function as administrative units out of which 5 of the 33 local government areas make up the state capital (12). Ibadan metropolis was the centre of administration of the old Western Region Nigeria since the days of the British colonial rule. The principal inhabitants of the state are the Yorubas. The city hosts quite a number of small, medium and large-scale industries involved in the production of food and beverages, clothing and textiles, chemicals and pharmaceuticals, confectionaries, leather-works and furniture, plastics, blocks etc (12). The study was carried out in 261 selected government-owned primary health facilities in Oyo State.

Primary healthcare workers directly involved in malaria case management in the selected government-owned primary health facilities in Oyo State were enrolled as participants in this study. This study employed a multistage sampling technique.

**Stage 1**: From the three senatorial districts in Oyo state, 8 LGAs were selected from each senatorial district using simple random sampling by balloting. **Stage 2**: Out of the selected LGAs in each senatorial district, 6 wards were selected using systematic sampling technique. **Stage 3**: From the 6 selected wards, using proportional allocation, a total of 261 PHCs was selected from the six wards. **Stage 4**: One healthcare worker providing malaria services (preferably the head or any health worker that was delegated by the head of malaria services) from each of the 261 selected PHCs was invited to participate in the study. All heads of primary health care workers who were not present in the facilities while data collection was ongoing were traced and enrolled. Visits were repeated at least thrice to ensure that all eligible frontline health workers were enrolled in the study.

A pretested standardized electronic structured questionnaire was used to collect information from the participants. The questionnaire consisted of 5 sections which included the socio-demographic characteristics of respondents, the knowledge of healthcare workers on malaria diagnosis and COVID-19, the practices of healthcare workers regarding malaria diagnosis, the practices of healthcare workers regarding COVID-19 differential diagnosis and challenges that healthcare workers face regarding malaria diagnosis. Data were coded, cleaned and analysed using SPSS version 25 software and MS-Excel. The dependent variables for this study were knowledge and practices of healthcare workers regarding malaria diagnosis and COVID-19 diagnosis while the independent variables were sociodemographic and socioeconomic characteristics. Means and SD were used to summarise the continuous variables while frequencies, proportions and charts were used to summarise the categorical variables.

Each knowledge and practice questions were scored and categorized into good and poor knowledge on malaria diagnosis, good and poor malaria-related diagnosis practices and good and poor ability to differentiate between malaria and COVID-19. Primary healthcare providers that scored 80% and above of the total possible scores at all levels (Knowledge and Practice) were dichotomized as good while those that scored below 80% were categorised as poor.

Chi-square tests were used at the bivariate analysis level to test the association between categorical outcomes. Binary logistics regression with respective odd ratios and confidence intervals were used at multivariate analysis to explore the relationship between the sociodemographic, socioeconomic characteristics and categorical outcome variables. The level of significance was set at p-value < 0.05.

Ethical approval for this study with approval number AD 13/479/4121 ^A^ was obtained from the Oyo State Ministry of Health, Ibadan prior to the conduct of the study. Permission and approval to conduct the study was also obtained from the Head of all the selected PHCs. In addition, the purpose of the study was explained to the participants and their written consents were obtained before the questionnaires were administered. Confidentiality and anonymity were ensured and the execution of the research was conducted according to the guidelines of World Medical Association (WMA) declaration of Helsinki regarding ethical guidelines and principles for conduct of research involving human subjects (13).

## Results

The study respondents totalled 261 primary health workers. The respondents ranged between 18 and 58 years were adults with a mean age of 44.3 years (±SD 8.5). The workers’ characteristics are presented in Table 1. Of the sample, 89.7% (234) were females and 92.7% (242) had post-secondary education. Less than one third (18.8%) were nurses, 90.0% (235) work at primary healthcare centres, 85.8% (224) had at least ten years of experience and 77.8% (203) earn above the national minimum wage (₦30,000 equivalent of $55). Majority of the respondents (96.2%) had participated in an Infection, Prevention and Control (IPC) training related to COVID-19 pandemic and 95.8% (250) were trained on malaria diagnosis and treatment. Only 12.3% (32) had their training on malaria diagnosis and treatment in the last five years and 75.9% (198) received online courses regarding COVID-19.

**Table 1:**
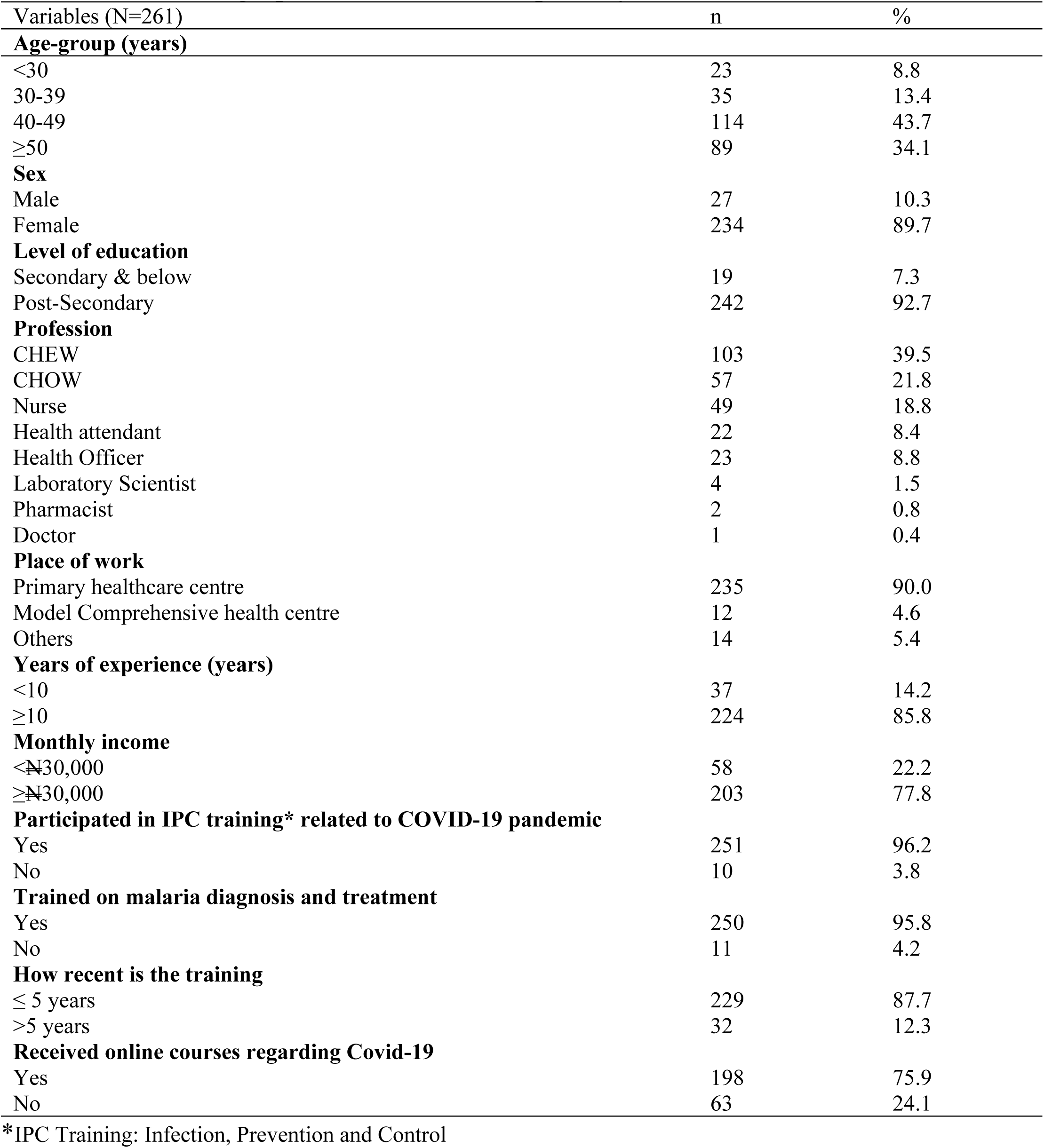
Socio-demographic characteristics of primary health care workers.

Figure 1 presented the level of knowledge of the primary healthcare workers on malaria and COVID-19. As seen in the chart more than two thirds (81.6%) of the respondents had good knowledge about malaria and 74.3% had good knowledge about COVID -19.

**Figure 1:**
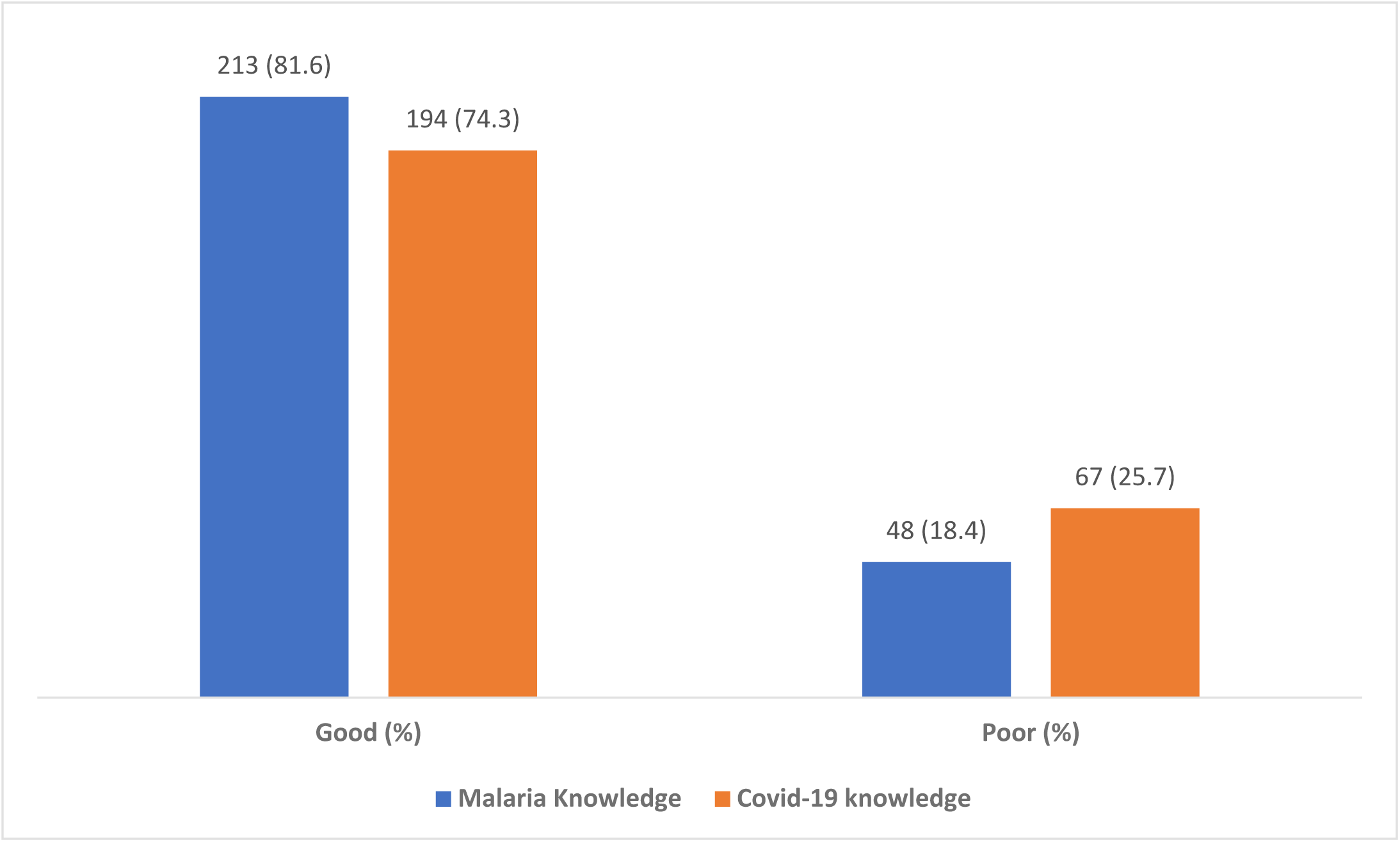
Overall knowledge score of primary healthcare workers on malaria and COVID-19.

Figure 2 highlighted the practice of primary healthcare workers on malaria diagnosis and COVID-19 differential practice. Relating to malaria diagnosis, majority (86.2%) of the respondents engaged in poor practices. The practices relating to COVID-19 differential diagnosis in 69.7% of the respondents were also poor.

**Figure 2:**
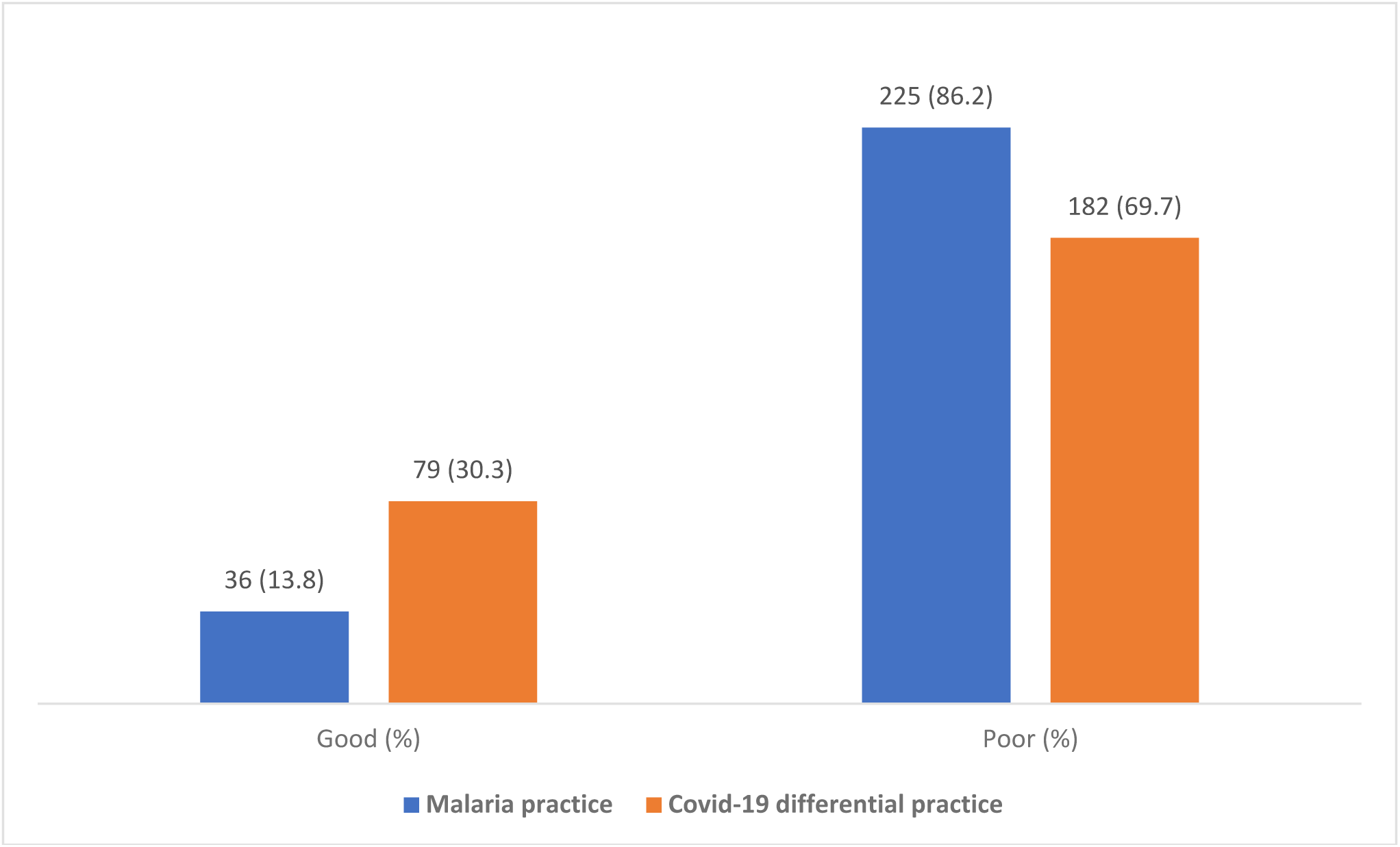
Overall practice score of primary healthcare workers on malaria and COVID-19 differential practice.

Figure 3 revealed the challenges commonly faced by the health workers with malaria diagnosis. Lack of adequate training on malaria diagnosis (27.8%) and unavailability of mRDT (18.4%) were the leading challenges with malaria diagnosis identified by healthcare workers.

**Figure 3:**
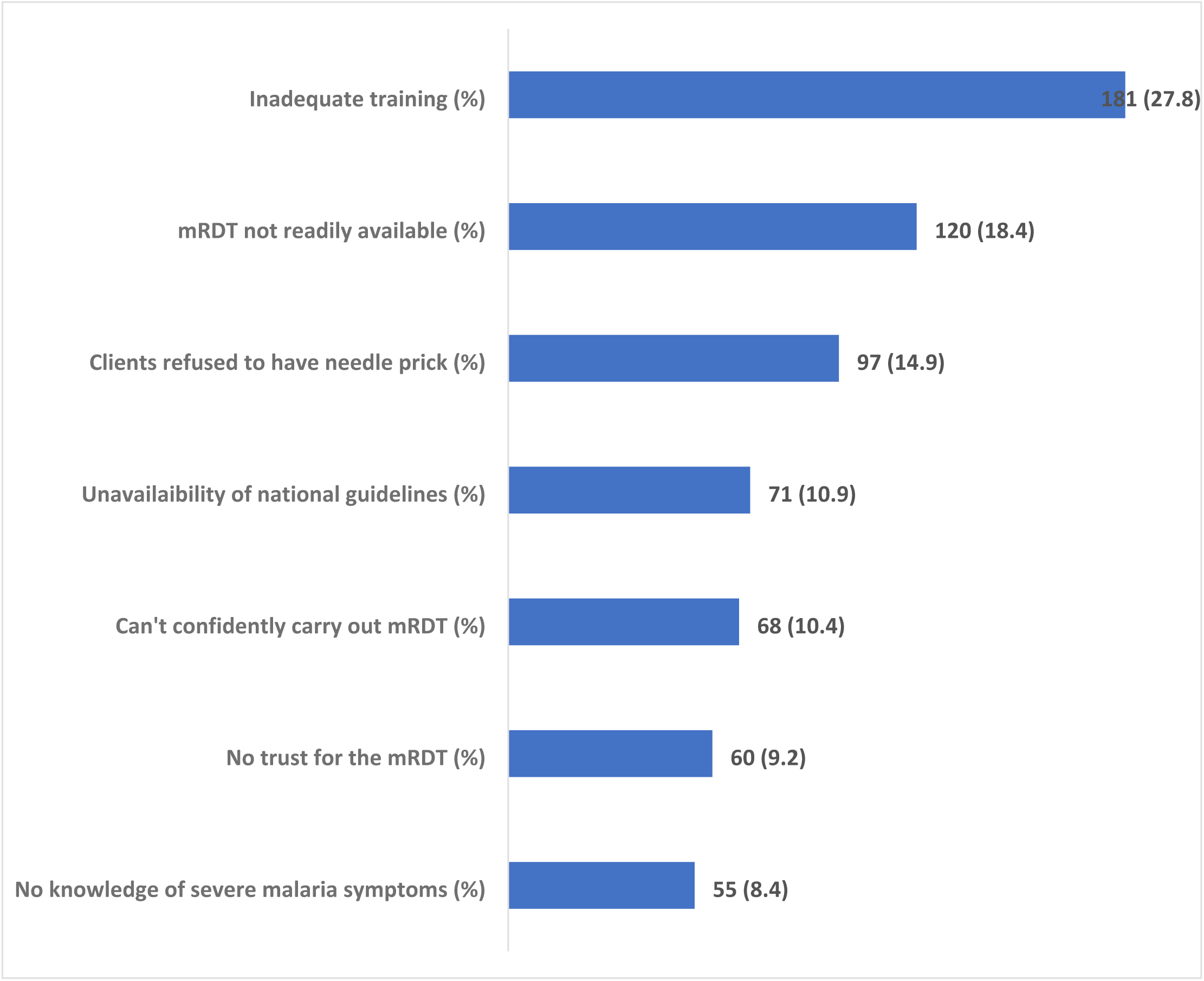
Challenges primary healthcare workers faced with malaria diagnosis.

Table 2. Variables that were significant at bivariate were subjected to multivariate analysis. Primary health care workers that received IPC training related to COVID-19 were 6 times more likely to have good knowledge on malaria compared to those that did not attend any related training (OR=5.936, 95%CI=1.265-27.857). Likewise, primary health care workers that received the training within the last 5 years were 3 times more likely to have good knowledge compared to those that received training more than 5 years ago (OR=3.296, 95%CI=1.367-7.946).

**Table 2:**
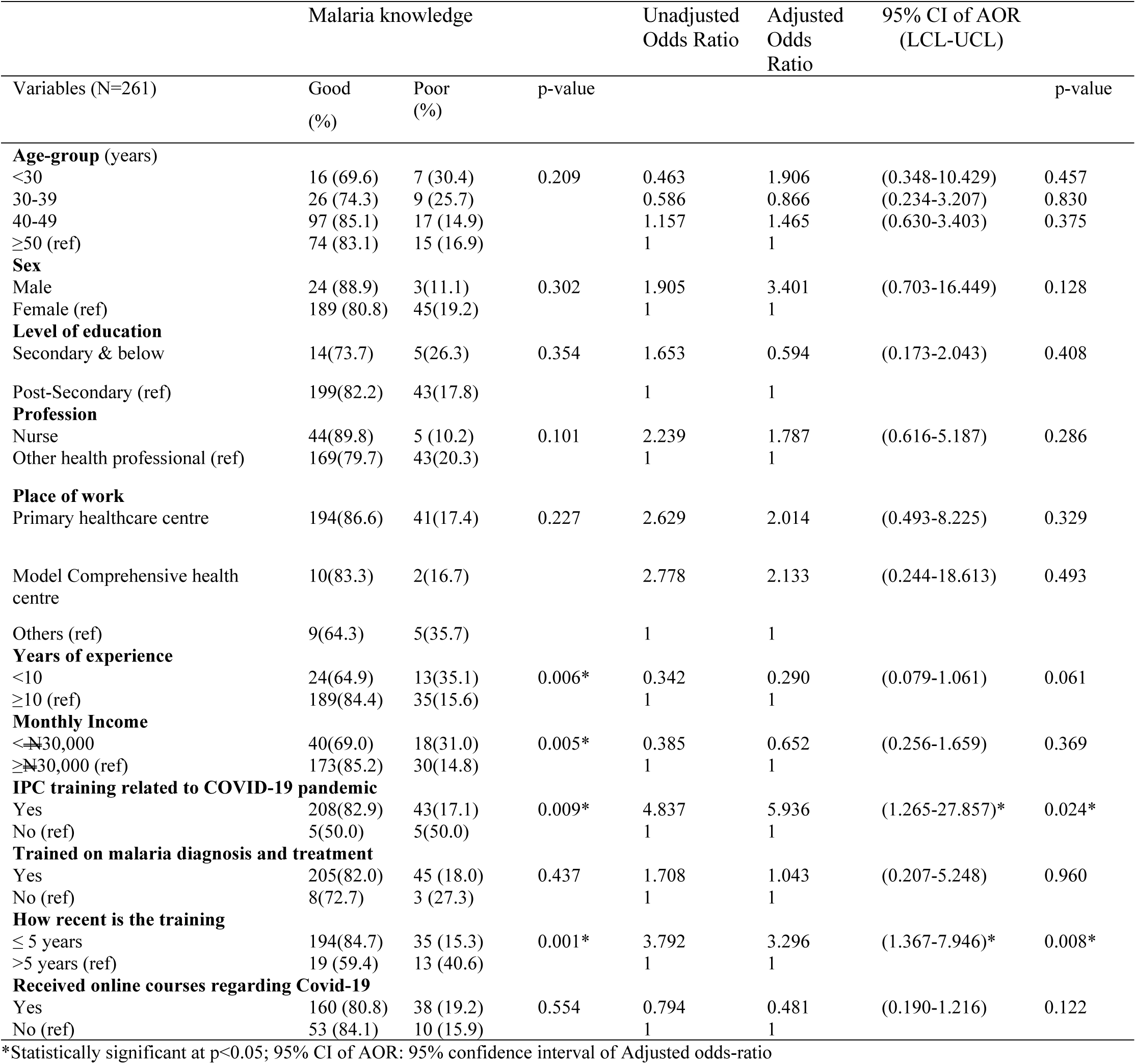
Factors influencing malaria knowledge of healthcare workers.

Table 3. Primary health workers that were nurses were 10 times more likely (OR=10.501, 95%CI=2.37-436.361) to have good knowledge on COVID-19 compared to the other health professionals. Primary health workers that work at the Model comprehensive health centre were also less likely (OR=0.113, 95%CI=0.015-0.859) to have good knowledge on COVID-19 compared to those working at the Primary health care centres.

**Table 3:**
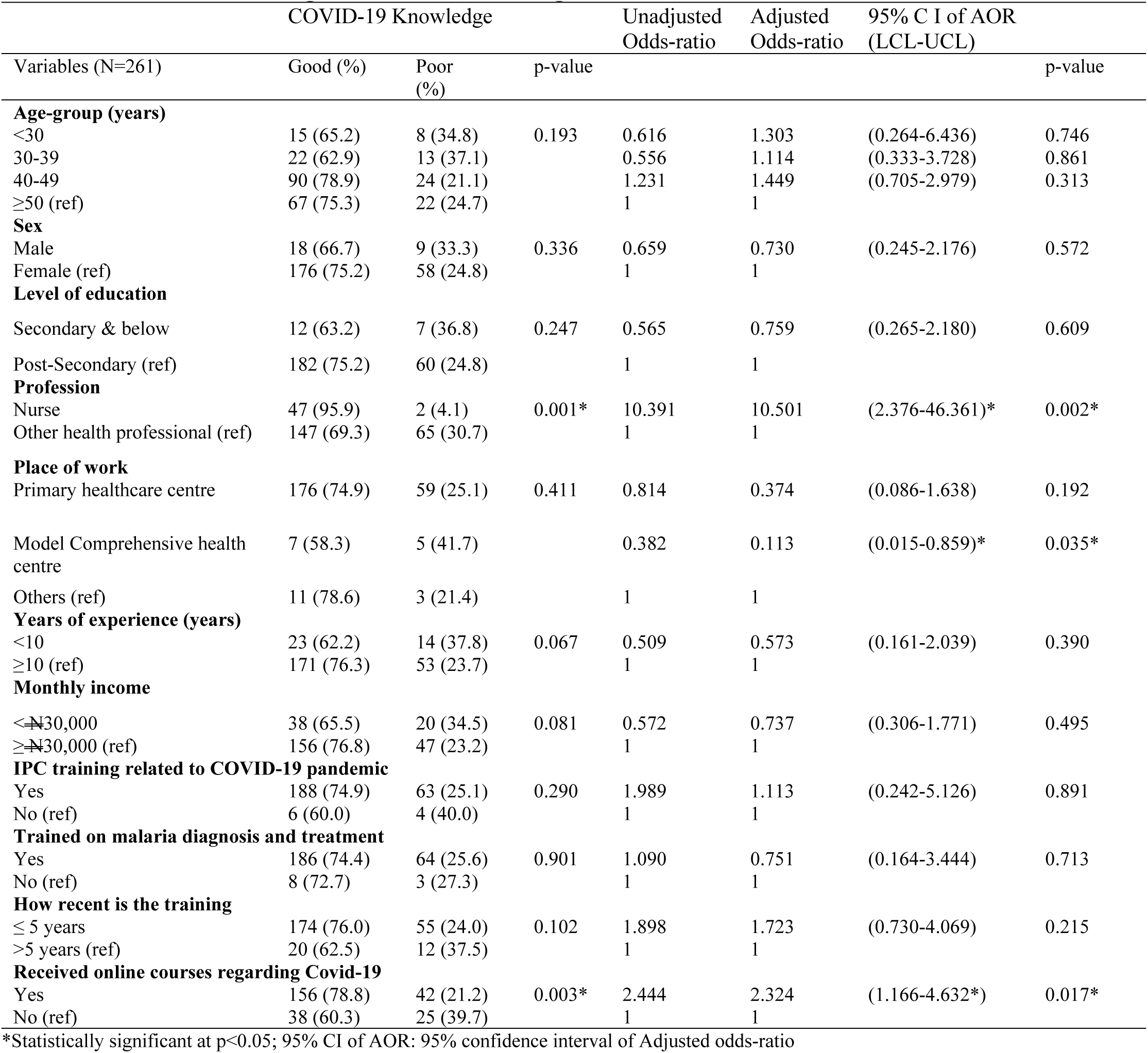
Factors influencing COVID-19 Knowledge of healthcare workers.

Table 4. Primary health workers within the age range of 30-39 were 4 times more likely (OR=4.388, CI=1.067-18.408) to adopt good practices relating to malaria diagnosis compared to aged 50 and above. Table 5. Primary health workers that earn less than the minimum wage were 4 times more likely (OR=3.989, CI=1.747-9.108) to have good COVID-19 differential diagnosis practices compared to those earning above minimum wage. Also, primary health workers who had training in the last 5 years were 3 times more likely (OR=3.491, CI=1.185-10.288) to have good COVID-19 differential practice compared to those that had training for more than 5 years. Primary health workers that received online courses regarding COVID-19 were 3 times more likely (OR=3.003, CI=1.340-6.734) to have good knowledge on COVID-19 differential diagnosis compared to those who did not.

**Table 4:**
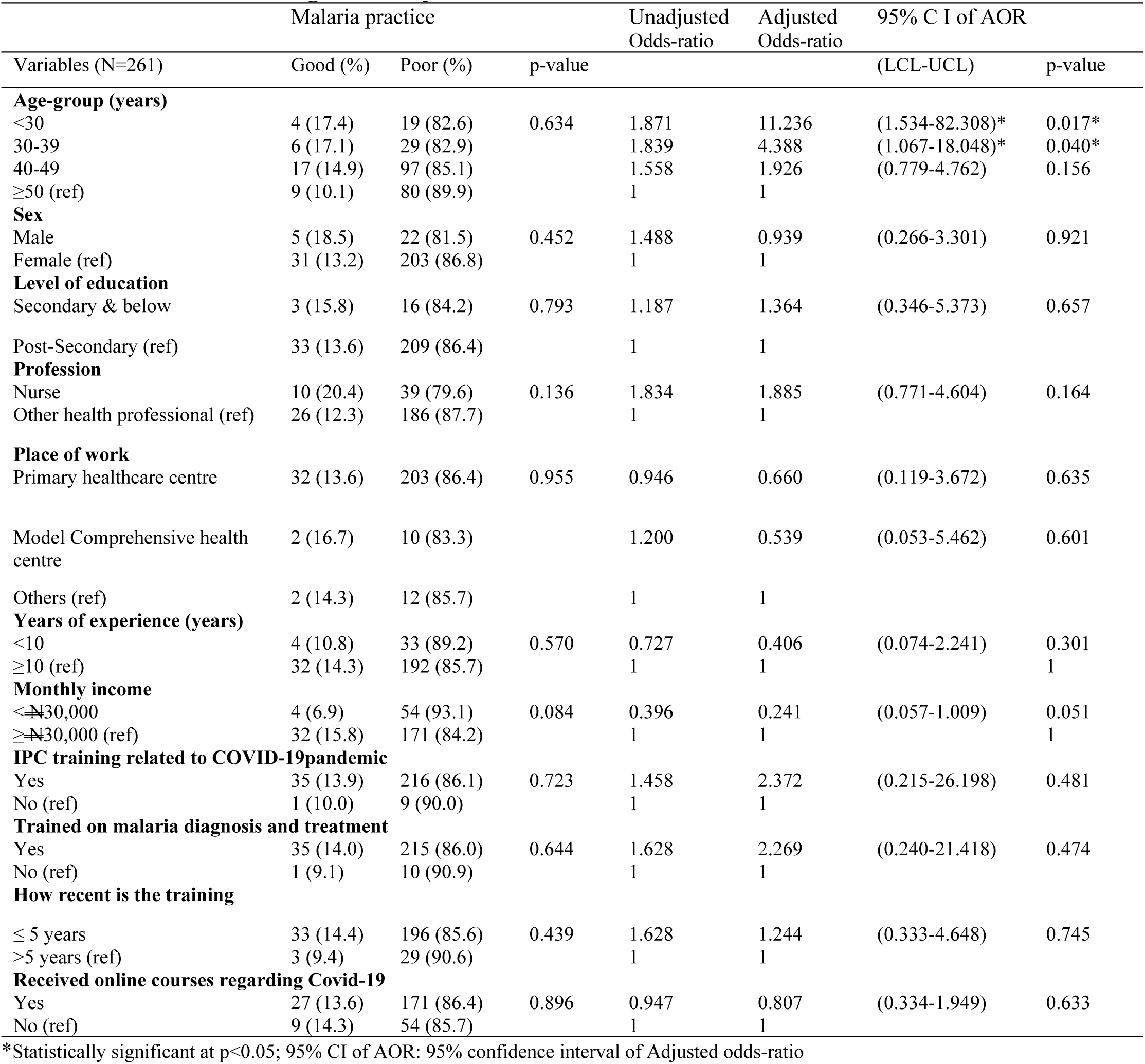
Factors influencing malaria practice of healthcare workers.

**Table 5:**
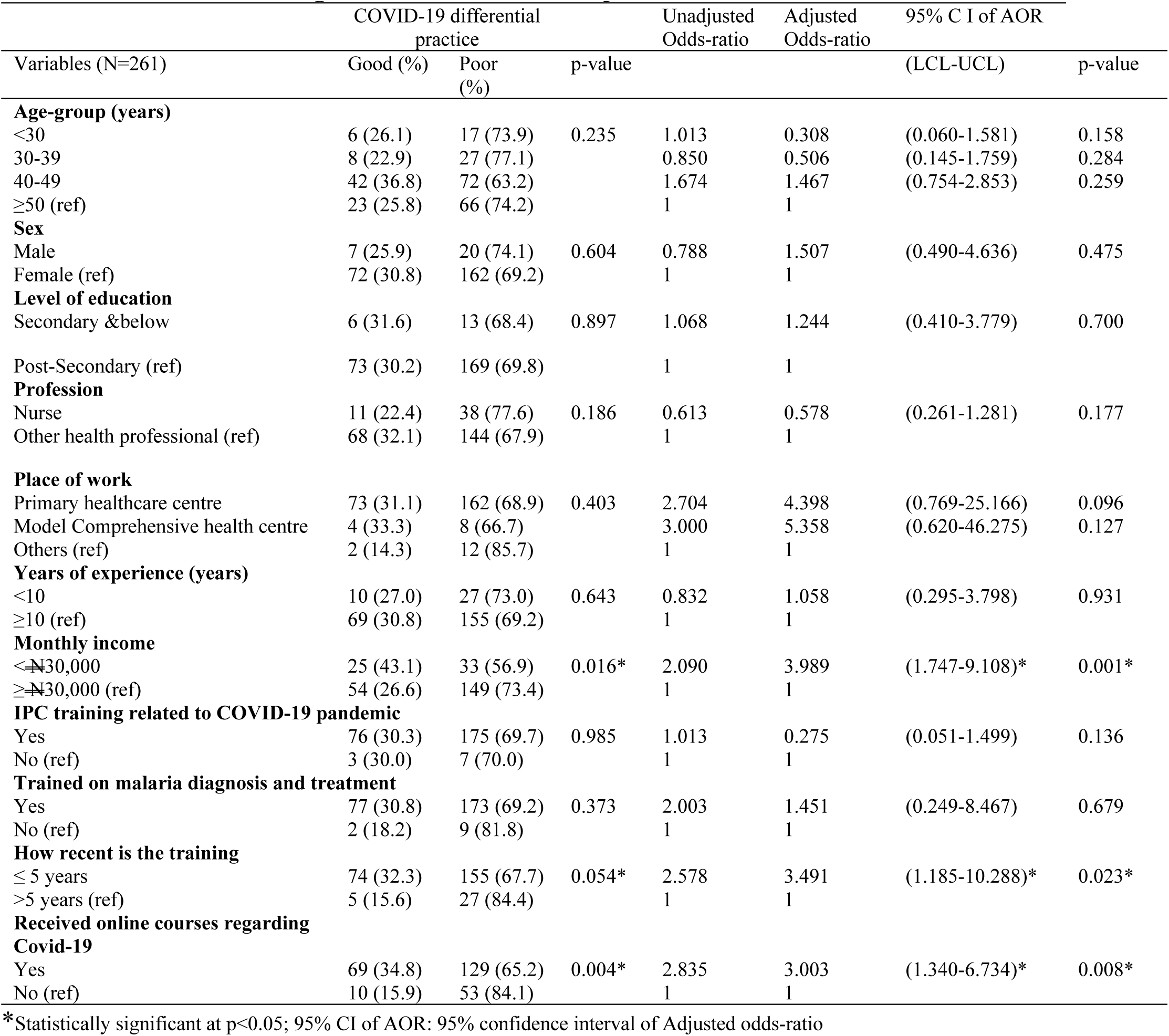
Factors influencing COVID-19 differential practice of healthcare workers.

## Discussion

Globally in 2018, about 228 million cases of malaria were reported with 405 thousand deaths, mainly coming from low-income countries. Sadly, Africa accounted for majority (93%) of cases, while six countries including Nigeria accounted for over half of the global burden of malaria (14).

Till date there is limited literature to understand co-infection with both diseases (6). Considering the limited resources available, the association between malaria and COVID-19 can be overwhelming in low- and middle-income countries. Nigeria battles with a very fragile healthcare system with poor healthcare infrastructures, unskilled and inadequate human resource and limited funding. Of the 424.4 million US dollars spent on malaria in 2016, only 19% was from the government (15). The resultant effect is availability of unqualified staff, little or no commodities to manage common ailments such as malaria at the primary healthcare level facilities.

### Access to laboratory diagnosis

The gold standard for malaria diagnosis is the malaria microscopy. Access to malaria microscopy especially in rural areas is limited, however due to donor funding, the use of the Rapid Diagnostic Test kits (RDT) has become a little popular in malaria diagnosis in Nigeria. On the other hand, suspected COVID-19 infection requires the use of a reverse transcriptase polymerase chain reaction (RT-PCR) in addition to clinical and radiological signs for diagnosis (16) in addition, rapid antigen test kits are largely unavailable for use in health facilities (17). The RT-PCR for the diagnosis of COVID-19 is not available at the primary healthcare or district levels in Nigeria. A patient presenting with symptoms suggestive of COVID-19 at this level, will leave the healthcare worker with her clinical acumen to suspect COVID-19 while she battles with determining whether it is malaria due to unavailability of malaria RDTs. The COVID-19 pandemic has no doubt also led to the disruption of supply chain for malaria RDTs as companies now focus more on the production of rapid tests for COVID-19 (14). The challenge remains availability of test kits and skills to perform the tests accurately resulting in a high rate of clinical diagnosis for malaria and unreliable test results respectively. This highlights the need to improve supply of these test kits to PHCs.

### Wrong diagnosis and wrong treatment

Our study showed that 75.9% and 95.8% of respondents had participated in trainings on COVID-19 and malaria diagnosis and treatment respectively resulting in good knowledge on both diseases. However, the good knowledge did not translate to practice, as 85.6% of respondents had poor malaria diagnosis practices and could not properly differentiate between malaria and COVID-19. This raises concerns as COVID-19 and malaria have many symptoms in common including fever, difficulty in breathing, fatigue and headache, this makes differential diagnosis difficult (18) especially in resource poor settings with limited access to laboratory diagnosis.

While it is necessary for frontline health care providers to have access to and be able to accurately carry out malaria RDT, patients with negative test results must be carefully managed (19) and should further raise the suspicion for COVID-19. Despite the high knowledge of malaria and COVID-19 noted in our study, 86.2% and 69.7% of respondents had poor diagnosis practices for malaria and COVID-19 respectively. Multiple organ failure in adults and respiratory distress in children are commonly seen in COVID-19 cases and remain major signs of severe malaria (11). Identifying and differentiating between cases presenting with such symptoms remains a huge challenge. The symptom overlap may lead to delay in treatment which may inadvertently affect the treatment outcome (20). Hence, there is a need to improve malaria diagnosis practices and COVID-19 differential diagnosis practices among primary healthcare workers to improve patient health outcomes.

### Training of healthcare providers and service integration

The primary healthcare remains the first line of reach to the communities (21) and several healthcare providers have been trained to manage malaria. To properly handle suspected COVID-19cases, the training on Infection, Prevention and Control (IPC) was rapidly scaled up to most healthcare providers (22), while online COVID-19 trainings were made available to providers (23). As revealed by this study, over 96% respondents have had recent training on COVID-19 related IPC while 75.9% has had online courses on COVID-19. These trainings are done in isolation of each other and does not converge to link the two diseases to create a better understanding. Linking these trainings will improve case identification, diagnosis and management for both illnesses. Provision of malaria supplies in laboratories where COVID-19 are carried out and vice versa will reduce missed opportunities for both diseases. Furthermore restructuring service provision, as was done for tuberculosis and HIV to provide a more collaborative approach will aid easy management of cases, this may include making available malaria RDTs where COVID-19 tests are done, further making diagnosis and case identification easy (24). The need for training, retraining and on-the-job mentoring of primary healthcare workers on the proper use of mRDT for malaria diagnosis to minimise clinical diagnosis and unreliable test results is vital to improve management of suspected cases at this level

## Limitations

Our study focused on the healthcare providers at the primary healthcare level thus limiting its applicability in terms of scope. The addition of secondary and tertiary healthcare facilities (with specialized COVID-19 diagnostic and malaria testing services) would have been desirable and is recommended for further studies.

## Conclusion

The study revealed a gap in the ability of healthcare workers to differentiate between COVID-19 and malaria cases. This is largely due to unavailability of rapid test kits for the diagnosis of both diseases, where they are available, 86.2% and 69.7% of respondents had poor diagnosis practices for malaria and COVID-19 respectively. The need for necessary healthcare packages including training, equipment and test kits should be provided to strengthen the diagnosis and management of febrile illnesses (19) Testing for COVID-19 and malaria should be harmonised and the screening tests for malaria and COVID-19 should be made available in healthcare facilities to reduce misdiagnosis and aid management of cases (11,25). The need to provide testing kits widely for healthcare providers and regular training to safely identify malaria cases is needed. Furthermore, adequate training for primary health workers to correctly differentiate between malaria and COVID-19 infections in the midst of the current pandemic (19) is also vital.

## Data Availability

The data for this research work has also been deposited in a public repository and is widely available for researchers (https://doi.org/10.6084/m9.figshare.19772920)

## Consent to publish

Not Applicable

## Availability of data and materials

The data that support the findings of this study is available on a public repository - **https://doi.org/10.6084/m9.figshare.19772920**

## Authors’ Contributions

EA conceptualized the study. EA and DB wrote the protocol, literature review and carried out the research. EA supervised the study. OA and OK performed the data analysis. EA, DB, OA and OK wrote the initial draft of the manuscript. TO revised the manuscript, provided technical and critical reviews on the improvement of the manuscript. All authors proof-read and approved the final manuscript.

## Acknowledgements

The authors thank all the medical directors and heads of department for granting the permission to conduct the interviews with the health workers. The authors also appreciate all the health workers that agreed to participate in this study. The efforts and time taken by the reviewers assigned to review this paper by the journal are also appreciated.

## List of abbreviations

COVID-19: Coronavirus disease 2019
HIV: Human Immunodeficiency Virus
IPC: Infection, Prevention and Control
LGA: local government areas
LMIC: low- and middle-income countries
LCL: lower confidence level
mRDT: Malaria Rapid Diagnostic Test
PHC: Primary Health Centres
RDT: Rapid Diagnostic Test
RT-PCR: Reverse Transcriptase Polymerase Chain Reaction
SARS-CoV-2: Severe Acute Respiratory Syndrome Coronavirus 2
SPSS: Statistical Packages for Social Sciences
UCL: Upper Confidence Level
WHO: World Health Organization
WMA: World Medical Association

